# Impact of COVID-19 Policies on the Post-pandemic Resurgence of Dengue Fever in Southeast Asia and Latin America

**DOI:** 10.64898/2026.01.07.26343578

**Authors:** Jiayi Chen

## Abstract

**Background:** Dengue fever, one of the most widespread mosquito-borne diseases, declined markedly during the early COVID-19 pandemic due to lockdowns and reduced mobility. However, the sharp resurgence in 2023 raised concerns about the long-term epidemiological effects of pandemic-era measures. This study examines the sustained impact of COVID-19 public health policies on dengue transmission across seven high-burden countries in Southeast Asia and Latin America.

**Methods:** Monthly dengue cases and COVID-19 policy indices from January 2020 to December 2023 were analyzed in seven countries with ≥2,000 dengue cases in 2020. Based on the Oxford COVID-19 Government Response Tracker (OxCGRT), countries were grouped as Strict COVID-Control (SCC: Singapore, China) or Less Strict COVID-Control (LSCC: Indonesia, Argentina, Brazil, Colombia, Mexico). Generalized linear models assessed lagged effects of the Stringency Index and policy sub-components, adjusting for climate and reporting bias.

**Findings:** Dengue cases declined by 41.3% (95% CI: 36.2–46.5; p < 0.001) in 2020 relative to pre-pandemic levels. In LSCC countries, public information and vaccination policies were significantly protective (β = −174,354; p < 0.001), reflecting indirect benefits of COVID-19 awareness infrastructure. In SCC countries, economic interventions showed mixed effects: debt relief reduced dengue risk (β = −0.783; p = 0.003), while income support increased it (β = 0.583; p = 0.003).

**Interpretation:** COVID-19 policies exerted lasting, heterogeneous effects on dengue transmission. Strengthening communication, vector control, and vaccination in LSCC settings, and maintaining vigilance and integrated surveillance in SCC settings, are crucial for sustainable dengue preparedness.

## 1. Introduction

Dengue fever remains one of the most significant vector-borne diseases globally, with an estimated 100–400 million infections annually, disproportionately affecting Southeast Asia and Latin America (WHO, 2025). Prior to COVID-19, the case number of dengue had been increasing steadily, driven by rapid urbanization, climate variability, and ineffective or inconsistently implemented vector control strategies (André et al., 2019). The onset of the COVID-19 pandemic profoundly disrupted the transmission dynamics of many infectious diseases, including influenza, respiratory syncytial virus (Groves et al., 2021), tuberculosis and malaria (Hogan et al., 2020). Previous studies documented sharp reductions in the case number of dengue during the early lockdown phases of the COVID-19 pandemic, primarily attributed to mobility restrictions and reduced human–mosquito contact (Araújo et al., 2024; Chen et al., 2022).

However, the incidence of dengue underwent a significant resurgence following the relaxation of COVID-19 policies, with 2023 witnessing the highest documented case number since 2000. In certain countries, such as Argentina, Brazil, and Singapore (Weng et al., 2025). While previous studies have described a short-term decline in dengue during COVID-19 (Chen et al., 2022), there remains limited evidence on how long-term policy stringency and the relaxation of these policies shaped post-pandemic resurgence across diverse governance contexts.

In the present study, we systematically evaluated the post-COVID burden associated with dengue fever across Southeast Asia and Latin America, two regions that account for the majority of global dengue cases and where COVID-19 policies were both diverse and stringently documented (Hale et al., 2021; WHO, 2025). We selected seven countries—China, Singapore, Argentina, Brazil, Colombia, Mexico, and Indonesia—on the basis of their high baseline dengue case number (≥ 2,000 reported cases in 2020) and availability of complete policy data. To allow for meaningful comparison, countries were selected into two groups using standardized criteria: Strict COVID Control (SCC) countries, defined as those with national lockdowns exceeding 50 consecutive days and an Oxford Stringency Index >80 during 2020–2022 (China and Singapore), and Less Strict COVID Control (LSCC) countries, which met neither threshold (Argentina, Brazil, Colombia, Mexico, and Indonesia). We constructed non-Bayesian regression models linking dengue case number to COVID-19 policy indices, with particular focus on economic support measures (e.g., income support, debt relief) and health-related interventions (e.g., public information campaigns, vaccination policy, and protective measures). This framework enabled us to evaluate how distinct policy domains differentially influenced the resurgence of dengue, and whether deviations from predicted trends were associated with shifts in physiological risk profiles, public awareness, or environmental conditions. By performing this research, we aimed to clarify the long-term impacts of COVID-19 on the epidemiology of dengue and to propose innovative and sustainable approaches to strengthen epidemic preparedness in the post-pandemic era. Our work contributes to the existing literature by providing one of the first multi-country, post-pandemic evaluations of dengue, with implications for policy design regarding how future pandemics may reshape the trajectory of endemic diseases.

## 2. Methods

### 2.1. Study design and participants

In this study, we focused on seven countries, three from Asia and four from South America, with at least 2,000 reported dengue cases in 2020, according to the OpenDengue database (https://opendengue.org/data.html) (OpenDengue, 2024). Monthly dengue case number for each country were extracted from the OpenDengue database between January 2020 and December 2023. Dengue case counts included both suspected and laboratory-confirmed cases. While reporting quality may vary between countries, reporting practices were assumed to be consistent within each country over time.

### 2.2. COVID-19 policy data

Monthly COVID-19-related policy indicators were retrieved from the Oxford Coronavirus Government Response Tracker (OxCGRT), as described by Hale et al. (2021). The OxCGRT Stringency Index was used as the benchmark variable for policy severity. In subsequent analyses, disaggregated policy variables, specifically the economic support and health system measures, were also extracted to assess the contribution of specific policies.

### 2.3. Climate adjustment

Since climate is known to be a major driver of dengue seasonality (Chen et al., 2022), we incorporated country-level climate controls into our regression model. Monthly mean temperature and seasonal rainfall patterns were extracted from publicly accessible ERA5 datasets (Hersbach et al., 2023). Where rainfall data were unavailable, a binary indicator for rainy versus dry season was applied. This adjustment aligned with that used in previous literature relating to the modeling of dengue (Chen et al., 2022).

### 2.4. Grouping method

Based on the stringency and duration of pandemic control policies, countries were selected into two groups. Countries were classified into the Strict COVID-Control (SCC) group if they met both of the following criteria: (1) national COVID-19 quarantine or lockdown measures extended for at least 50 consecutive days, and (2) a mean OxCGRT Stringency Index value > 80 for the 2020–2022 period. Using these criteria, China and Singapore were categorized as SCC countries. All other countries (Indonesia, Argentina, Brazil, Colombia, and Mexico) were categorized into the Less Strict COVID-Control (LSCC) group. This grouping method aligned with previous work (Chen et al., 2022; Hale et al., 2021; Islam et al., 2021) and facilitated the comparison of long-term trends in dengue transmission under different public health governance models.

### 2.5. Statistical analysis

We applied a generalized linear model (GLM) to investigate the association between COVID-19 policy measures and the monthly dengue case number. The primary equation is given in Eq. (1).

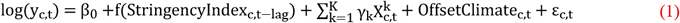

In Eq. (1), y_c,t_ represents the monthly case number of dengue for country c at time t, StringencyIndex_c,t−lag_ represents the OxCGRT stringency index for the prior month; f(·) represents a natural cubixc spline function; 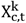 represent covariates, including temperature, rainfall (or seasonal dummies), and monthly fixed effects; OffsetClimate_c,t_ represents the climate data collected to offset the impact of confounding variables; and ε_c,t_ represents the model error term.

Models were fitted separately for SCC and LSCC groups to capture potential differences in policy-disease dynamics. All regressions incorporated a one-month lag to reflect typical mosquito development and disease incubation cycles, consistent with previous modeling frameworks (Brauner et al., 2021).

Other than GLM, we also conducted principal component analysis (PCA) and cluster analysis on the data for closer analysis. PCA was essential for identifying the key policy dimensions driving variation in dengue dynamics across countries, reducing complex, correlated variables into a few interpretable components that explained over 90% of total variance. Cluster analysis further complemented this by uncovering distinct dengue case patterns within each policy group, revealing how differing combinations of interventions corresponded to markedly different disease burdens.

Model estimates were computed using EViews version 13 software (IHS Markit, 2024).

### 2.6. Under-reporting considerations

We acknowledge that the under-reporting of dengue may have occurred during the COVID-19 pandemic due to healthcare system strain and testing disruptions. Previous studies, including Islam et al. (2021), demonstrated that care-seeking behavior declined during the early phases of the pandemic. While this may have impacted absolute counts, we assumed that trends within each country were consistent over time, given that the same national surveillance infrastructure was applied across all months in the designated period.

## 3. Results

Our analysis revealed distinct policy-dengue relationships between SCC and LSCC countries.

We analyzed dengue case number in countries with less strict COVID control (LSCC: Argentina, Brazil, Colombia, Mexico, Indonesia) and strict COVID control (SCC: Singapore, China) using generalized linear model. In the LSCC group, public information campaigns (H1) show significant negative association with dengue case number (coefficient = −174,354, *p* < 0.001). Negative associations were also significant for facial coverings (H6; *p* = 0.022) and vaccination policy (H7; *p* = 0.038), whereas income support (E1) showed a marginally positive association (*p* = 0.095). In the SCC group, economic policies show highest correlation: income support (E1) was positively associated with case number (coefficient = 0.583; *p* = 0.003), while debt relief (E2) demonstrated a significant negative association (coefficient = −0.783; *p* = 0.003).

Principal component analysis (PCA) revealed that three components captured over 90% of variance in both groups. In LSCC countries, the first component was primarily driven by H1 and H7, whereas in SCC countries, variance was shaped by E1, E2, and elder protection (H8). Cluster analysis identified three distinct disease patterns per group, with the highest cluster averaging 880,880 cases per month in LSCC and 2,907 cases per month in SCC.

Model diagnostics indicated robust performance. Residuals in SCC models passed homoskedasticity and autocorrelation checks, while LSCC models exhibited mild heteroskedasticity but stable variance overall. Statistical power was consistently high (≥ 0.85), with large effect sizes (Cohen’s f^2^ ≥ 0.60). All tests were two-sided, with significance set at *p* ≤ 0.05.

## 4. Discussion

This study systematically evaluated the long-term effects of COVID-19 policy measures on the case number of dengue across seven endemic countries, addressing a key gap in the existing literature. Prior studies have primarily focused on short-term declines in dengue during the early phases of the pandemic (Chen et al., 2022;

Araújo et al., 2024), but few have assessed how policy stringency and duration influenced post-pandemic resurgence. By grouping countries into Strict COVID Control (SCC) and Less Strict COVID Control (LSCC), we demonstrated how variations in policy enforcement and baseline public health capacity shaped divergent epidemiological trajectories. These findings are significant because they highlight the importance of tailoring dengue control strategies to policy environments, providing a framework for adaptive preparedness in future pandemics.

Our analyses demonstrated that the impact of COVID-19 policy measures on dengue varied considerably between SCC and LSCC countries and that these variations were shaped by differences in baseline awareness, policy enforcement, and public health infrastructure. In LSCC settings, where dengue burden remained high and governance was more fragmented, public information campaigns (H1) and vaccination policies (H7) were strongly protective (*p* < 0.01). These findings suggest that targeted, low-cost communication strategies, combined with preventive interventions such as vaccination, can significantly mitigate dengue risk in less centralized health systems. In contrast, in SCC countries, economic support policies (E1) and debt relief (E2) showed statistically significant associations with dengue case number, with E1 positively associated (*p* = 0.003) and E2 negatively associated (*p* = 0.003). This indicates that while economic support correlated with higher reported case number, debt relief correlated with lower case number, reflecting opposing behavioral and socioeconomic influences on dengue risk.

When compared with previous research, our results were both consistent and divergent. Chen et al. (2022) documented an abrupt decline in dengue case number during the first year of the pandemic, particularly in high-stringency contexts, supporting our finding that strict policies suppressed transmission by reducing mosquito– human contact opportunities. Islam et al. (2021) similarly reported that reduced health-seeking behavior contributed to fewer reported dengue cases, thus aligning with our interpretation that under-reporting played a role in shaping observed case count trends during the pandemic. However, our results differ from studies in Brazil and Bangladesh (Araújo et al., 2024), which emphasized climatic rebound as the primary driver of post-2022 dengue resurgence. This divergence may be explained by regional variation in the stringency, duration, and enforcement of COVID-19 policies, underscoring the importance of group comparative analysis.

The methodological contribution of this study lies in demonstrating the usefulness of grouping countries by COVID-19 policy stringency and duration when investigating long-term dengue case number. Unlike earlier studies that focused primarily on short-term declines in 2020, our generalized linear regression framework captured sustained changes through 2023 and showed that policy effects persisted beyond the immediate lockdown phase. This highlights the context dependency of policy effectiveness: strong baseline surveillance and awareness in SCC countries limited the incremental benefit of additional information campaigns, whereas in LSCC settings, such interventions retained substantial marginal value.

Collectively, our findings suggest that future public health strategies should be tailored to baseline system capacity. In SCC countries, refining economic support policies to maintain household-level vigilance without inadvertently increasing exposure risk may be crucial. In LSCC countries, institutionalizing information campaigns and vaccination as long-term, low-cost preventive measures could provide sustainable benefits. More broadly, these findings emphasize how pandemic-era policy legacies continue to shape the epidemiology of endemic diseases and reinforce the need for adaptive, context-sensitive preparedness strategies.

Despite the strengths of this study, several limitations must be acknowledged. First, reliance on publicly available OxCGRT policy indices, while standardized, may not have fully captured on-the-ground policy enforcement or compliance variability across regions. Second, dengue case reporting may have been subject to under-reporting, particularly in overstretched health systems during the pandemic, although inclusion of countries such as Indonesia was intended to mitigate this concern. Third, climate variables were excluded from our final models due to data accessibility constraints and our focus on policy impacts. Future research should consider integrating temperature and precipitation data using ERA5 or comparable proxies. Finally, the generalizability of findings is limited by the sample size and regional scope, restricting extrapolation to non-endemic or low-transmission settings.

## 5. Conclusion

This multi-country analysis demonstrated that COVID-19 control policies exerted collateral impact on the case number of dengue, with effects mediated by governance strictness and public awareness. In LSCC countries, health communication and vaccination policy proved most protective, while in SCC countries, economic measures shaped the resurgence of dengue. To prevent the post-pandemic resurgence of endemic diseases, LSCC countries should institutionalize scalable information and vaccination campaigns, while SCC countries should strengthen localized and data-driven interventions. Future work should integrate fine-grained climate and mobility data. This study contributes to post-pandemic epidemiology by offering one of the first comparative assessments of COVID-19 policy spillovers on dengue, providing evidence-based guidance for epidemic preparedness.

## Acknowledgements

Not applicable.

## Funding

This research did not receive any specific grant from funding agencies in the public, commercial, or not-for-profit sectors.

## Declaration of competing interest

The authors declare that they have no known competing financial interests or personal relationships that could have appeared to influence the work reported in this paper.

## Data availability

Data will be made available on request.

## Figure captions

**Fig. 1.**
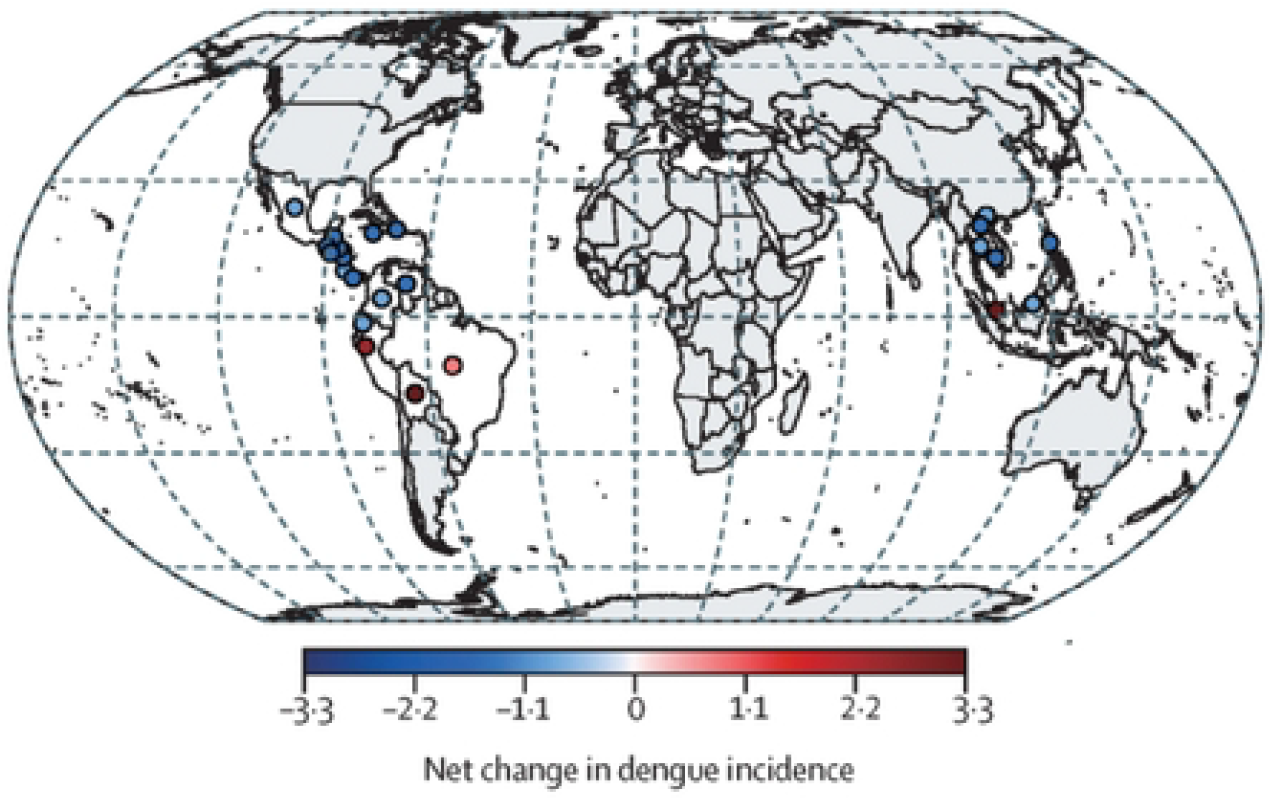
Changes in the global incidence of dengue during the COVID-19 pandemic (Chen et al., 2022)

**Fig. 2.**
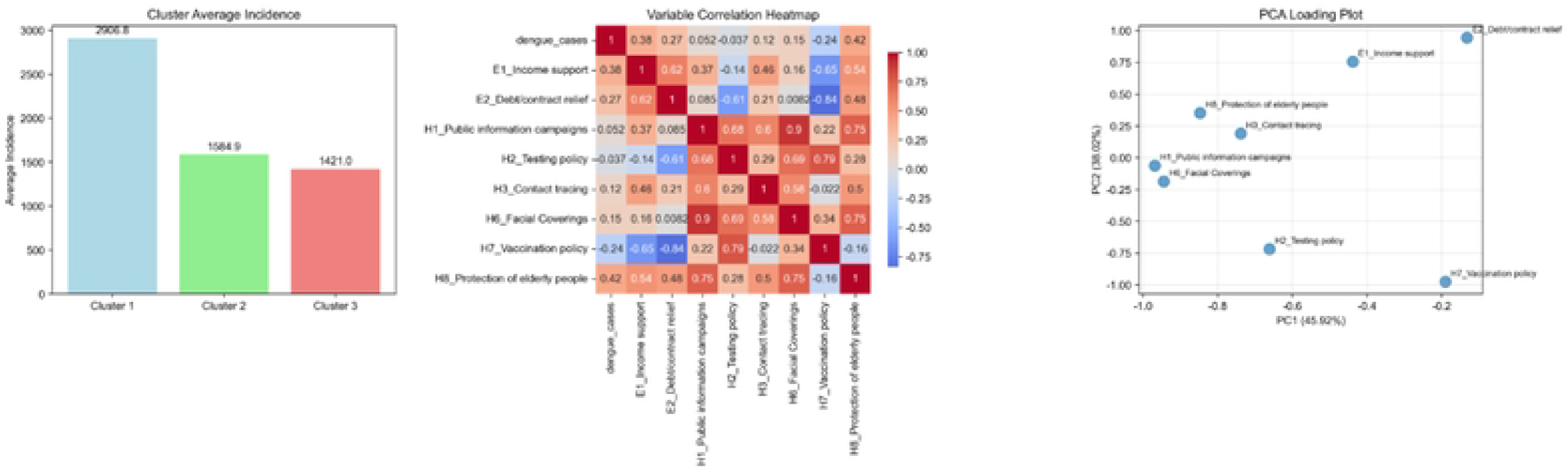
Correlation analysis for covariates in Strict COVID Control (SCC) countries

**Fig. 3.**
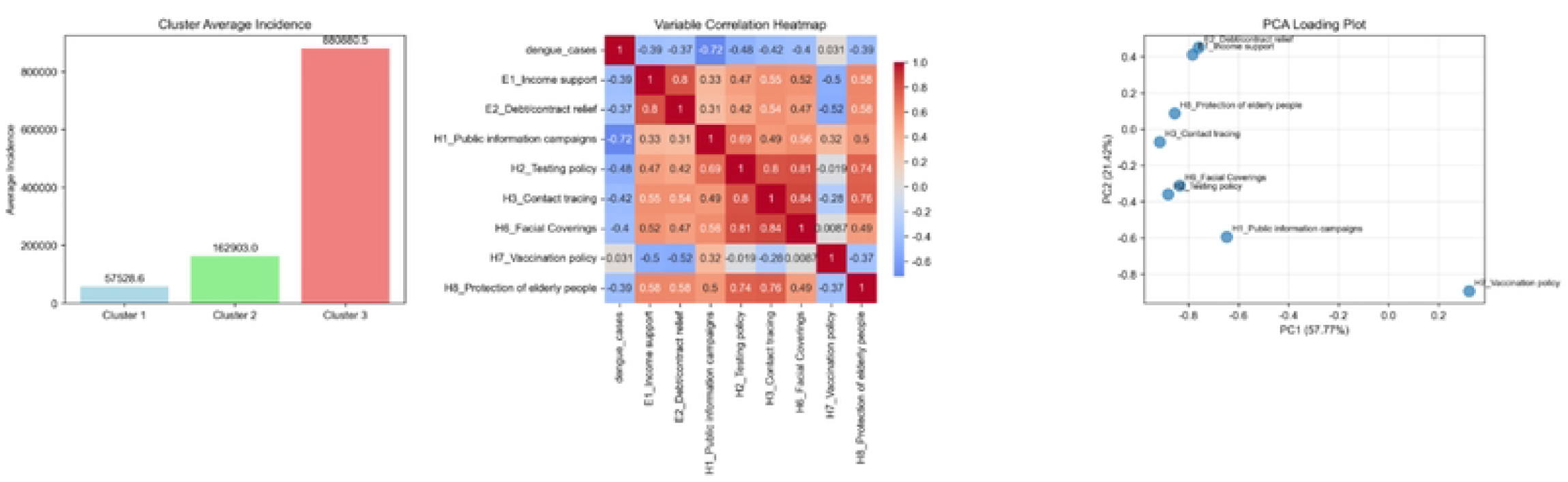
Correlation analysis for Less Strict COVID Control (LSCC) countries

## Notes

### Competing Interest Statement

The authors have declared no competing interest.

### Funding Statement

The author(s) received no specific funding for this work.

### Author Declarations

No IRB/oversight body is included in the study.

